# Acute associations between ambient air pollution and risks of preterm and early-term births: results from 8 states in the United States

**DOI:** 10.64898/2026.07.19.26358434

**Authors:** Xiaping Zheng, Amy Fitch, Joshua L. Warren, Hua Hao, Matthew J. Strickland, Andrew J. Newman, Lyndsey A. Darrow, Howard H. Chang

## Abstract

Ambient air pollution during pregnancy has been linked to adverse pregnancy outcomes, but evidence of acute exposure and shorter gestation length remains inconsistent. We examined the association between ambient air pollution and preterm (28-36 weeks) or early-term (37-38 weeks) births. Daily concentrations of 12 bias-corrected model-derived pollutants were linked to vital records of 1,085,162 preterm and 3,901,185 early-term singleton live births (2005-2017) across eight U.S. states. Under a time-stratified case-crossover design, odds ratios (OR) were estimated, adjusting for risks among ongoing pregnancies, meteorology, time trends and federal holidays. We estimated cumulative associations up to a 6-day lag using distributed lag models. Risk estimates per interquartile range increase were pooled across states using inverse-variance weighting. We observed positive associations between 0-2 day cumulative exposure to several air pollutants and early-term births, including NO_2_ (OR: 1.0023, 95% CI:1.0010, 1.0037 per 7.1 μg/m^3^), PM_2.5_ (OR=1.0022, 95% CI: 1.0006, 1.0038 per 4.6 μg/m^3^), PM_2.5_ organic carbon (OR= 1.0026, 95% CI: 1.0013, 1.0039 per 1.7 μg/m^3^) and PM_2.5_ elemental carbon (OR=1.0025, 95% CI: 1.0014, 1.0035 per 0.26 μg/m^3^). Associations with preterm birth were mostly null. Our findings suggest positive associations between short-term air pollution exposure, including PM and major PM_2.5_ components, and risks of early-term birth.

## 1. Introduction

Preterm birth (PTB), defined as delivery before 37 completed weeks of gestation, remains a major public health challenge globally and in the United States.^1^ It is a leading cause of neonatal morbidity and mortality^2^ and contributes substantially to long-term adverse health outcomes,^3^ including neurodevelopmental impairment, chronic respiratory disease, and cardiometabolic disorders later in life. Despite advances in obstetric and neonatal care, PTB continues to impose a significant healthcare and societal burden.^4^ In addition to PTB, early-term birth (ETB; 37–38 weeks of gestation) has emerged as an outcome of increasing clinical importance.^5^ Although traditionally considered within the normal range, ETB has been associated with elevated risks of respiratory complications, neonatal intensive care unit admission, and developmental delays compared to full-term births.^5^ Together, PTB and ETB represent a substantial burden on maternal and child health, underscoring the need to identify modifiable environmental risk factors that may contribute to reduced gestational length.

Ambient air pollution has been widely studied as a potential environmental determinant of adverse birth outcomes.^6–8^ A large body of epidemiologic evidence suggests that exposure to pollutants such as fine particulate matter (PM_2.5_), nitrogen dioxide (NO ), ozone (O ), and carbon monoxide (CO) during pregnancy is associated with increased risk of PTB.^9^ However, most prior studies and meta-analyses have focused on average or cumulative exposures across pregnancy or specific trimesters,^9^ with relatively consistent but modest associations. In contrast, evidence for short-term (acute) exposure immediately preceding delivery remains limited and mixed. A smaller number of studies using time-series, case-crossover, or time-to-event designs have examined exposures within days prior to birth, hypothesizing that transient increases in air pollution may act as triggers for labor.^10,11^ Large population-based studies have reported positive associations for ozone exposure in the days leading to delivery, while finding null or inconsistent associations for other pollutants such as PM_2.5_, NO_2_, and CO.^11,12^ These inconsistencies may reflect differences in exposure assessment, pollutant mixtures, study design, and study populations. Importantly, understanding acute exposure effects is critical for informing regulatory standards, real-time air quality warnings, and targeted interventions aimed at reducing exposure during high-risk periods of pregnancy.

A second major research gap concerns the relative toxicity and sources of air pollution most relevant to PTB and ETB. Air pollution is a complex mixture derived from diverse sources, including traffic emissions, industrial activities, and increasingly, wildfires.^13^ Emerging evidence suggests that source-specific exposures may have distinct health effects. For example, traffic-related air pollution has been linked to adverse birth outcomes, yet recent comprehensive evaluations indicate that evidence for its association with PTB remains suggestive but of low confidence, highlighting ongoing uncertainty in the field.^14^ At the same time, wildfire smoke has become an increasingly important contributor to ambient PM_2.5_, particularly in the United States, where it is eroding decades of improvements in air quality.^15^ Recent large-scale studies have reported positive associations between prenatal exposure to wildfire-related PM_2.5_ and increased risk of PTB, with evidence of critical exposure windows during pregnancy.^15^ Additional studies further suggest that wildfire smoke exposure and other combustion-related pollutants may influence PTB risk through pathways such as inflammation, oxidative stress, and placental dysfunction.^16^ Despite these advances, most prior studies have examined a limited number of pollutants, making it difficult to disentangle source-specific effects or identify the most harmful components of air pollution mixtures.

To address these knowledge gaps, we conducted a large multi-state study to estimate short-term associations between ambient air pollution and risks of both PTB and ETB across diverse populations in the United States. Using vital records from eight states between 2005 and 2017, we applied a time-stratified case-crossover design to assess acute exposures in the days preceding delivery. This study makes several key contributions. First, it represents one of the largest investigations to date in the United States examining short-term air pollution exposure and gestational timing, encompassing geographically and demographically diverse populations. Second, we extend prior research by explicitly examining ETB, an outcome that has been relatively understudied despite its clinical and public health significance. Third, we incorporate a comprehensive set of twelve pollutants, including major constituents of PM_2.5_ that may serve as proxies for specific sources, such as traffic and combustion-related emissions. By addressing both the timing of exposure and the complexity of pollutant mixtures, this study aims to provide more robust evidence on the role of acute air pollution exposure in shaping birth timing and to inform future research, regulatory policies, and targeted public health interventions.

## 2. Data and Methods

### 2.1. Birth Records and Environmental Exposure Data

Live birth records were obtained from eight states during 2005-2017: California, Florida, Georgia, Kansas, Nevada, New Jersey, North Carolina (only up to 2015), and Oregon. This study included singleton births with gestational age between 28-36 weeks (PTB) and 37-38 weeks (ETB). Births were excluded if maternal residential address was missing, out-of-state, or could not be matched to air pollution and meteorology data. We further excluded PTBs born after August 31 and ETBs born after October 31 in the last year of data collection. This addresses the fixed-cohort bias^17^ occurring near the end of the study period, where pregnancies with shorter gestational lengths were over-represented given the same conception date.

We utilized a daily data fusion product for 12 pollutants: CO, NO_x_, NO_2_, SO_2_, O_3_, PM_2.5_, PM_10_, and PM_2.5_ constituent SO ^2−^, NO ^−^, NH ^+^, elemental carbon (EC) and organic carbon (OC), available at 12-km spatial resolution. Pollution concentrations were estimated by bias-correcting simulations from the Community Multiscale Air Quality (CMAQ) model with monitoring measurements in the US Environmental Protection Agency’s Air Quality System database and land-use variables.^18^

We also acquired daily maximum and minimum temperatures at 1 km resolution from the High-resolution Urban Meteorology for Impacts Dataset (HUMID).^19^ HUMID has the advantage of improved characterization of spatial temperature variability in urban areas and across urban-rural divides, achieved through the integration of an energy balance urban canopy model into a physics-based land surface modeling system. It is bias-corrected with available station observations.

Both daily air pollution and daily mean temperature were spatially averaged to the 2010 ZIP Code Tabulation Areas (ZCTA) published by the US Census Bureau. For New Jersey and North Carolina, residential addresses were geocoded and linked to ZCTA polygons. For other states, maternal residential ZIP code was used, and births with ZIP codes that did not match with a ZCTA were excluded.

### 2.2. Statistical Analysis

We used a time-stratified case-crossover design to estimate short-term associations between air pollution exposure and risks of PTB and ETB. In the case-crossover design, for each PTB or ETB, we created 3 or 4 matched reference days defined by same day of week, month and year, which inherently controls for time-invariant confounders, such as maternal demographic characteristics and chronic comorbid conditions.

PTB and ETB were analyzed separately via conditional logistic regression. The primary exposure window of interest is the week prior to an event or a reference day. The models included daily air pollution exposures from lag 0 to lag 6. The distributed non-linear lag framework (DLNM) was used to allow for non-linear lag effects (natural cubic spline with 3 df, single knot placed at lag 3), assuming a linear pollution association at each lag. From the DLNM model, we extracted odds ratios associated with same-day exposure (lag 0), cumulative exposure over the last 3 days (lag 0-2), or over last 7 days (lag 0-6) association. Odds ratios were expressed per interquartile range (IQR) increase in pollutant concentration. For each pollutant, the IQR was calculated as the median of ZIP code–specific IQRs, each derived from daily concentrations during the study period.

For time-varying confounders, we included a binary federal holiday indicator and daily average temperature over the same exposure period (lag 0-6 days), assuming non-linear lag-specific effects (natural cubic splines with 6 degrees of freedom) that are unconstrained across lags.

Time-varying unmeasured confounding was accounted for by including natural cubic splines indexed by the birthdate’s day of year (i.e., 1, 2, …, 366) with 4 degrees of freedom to capture additional within-month trends smoothly. The above modeling choices mirror prior case-crossover studies on air pollution and morbidity (e.g., hospitalization and emergency department visits).

We further adjusted for within-window changes in PTB/ETB risk due to conception seasonality and digit preference in last menstrual period reporting. Specifically, we included a variable *W_i_*, defined as the ZCTA-level daily probability of case occurrence (28-36 weeks for PTB and 37-38 weeks for ETB) on day *i*. For PTB, 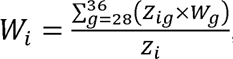 where is number of ongoing pregnancies at risk of birth at gestational week *g* on day *i*; *W_g_* is the conditional probability of birth at gestational week *g*, given that the gestational week is at least *g* weeks; and *Z_i_* is the total number of ongoing pregnancies at risk of PTB on day *i* (i.e., 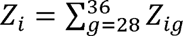). Similarly, *W_i_* was defined for ETB as 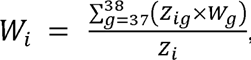 where 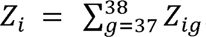.

State-specific estimates were obtained by fitting models within each state and then combined via inverse-variance weighting. Several sensitivity analyses were conducted to examine the robustness of our results. First, we considered a longer exposure windows of 2 weeks (lag 0-14). Second, we varied the birthdate day of year’s natural cubic splines to 5, 6, 7 and 8 degrees of freedom.

We assessed effect modification through stratified analysis by maternal age groups (< 25, 25-34, > 34)^1^, race/ethnicity (non-Hispanic White, non-Hispanic Black, Hispanic, Other), and maternal education (less than high school, high school or equivalent, and more than high school). These modifiers were chosen given the availability of maternal characteristic variables and their known associations with risks of PTB and ETB.

## 3. Results

The study included a total of 4,986,347 singleton live births, with 22% being PTB and 78% being ETB. Births from California represented 41.9% of the total sample, followed by Florida (19.8%) and Georgia (12.5%). State-specific sample sizes are given in Supplemental eTable 1. Maternal characteristics of PTB and ETB are summarized in Table 1. The majority of cases were born of pregnant individuals between the ages of 25 to 34, Hispanic or non-Hispanic White, and of more than high school education. Table 2 gives summary statistics of air pollution concentrations on the event or reference days. Average air pollution concentrations and variability were similar across PTB and ETB.

**Table 1.** Maternal demographics of preterm and early-term births from 2005-2017 in California, Florida, Georgia, Kansas, Nevada, New Jersey, North Carolina (2005-2015), and Oregon.

|  | **PTB** | **ETB** |
| --- | --- | --- |
| Total | 1,085,162 | 3,901,185 |
| Age |  |  |
| < 25 | 342,090 (31.5%) | 1,152,221 (29.5%) |
| 25-34 | 535,175 (49.3%) | 2,048,108 (52.5%) |
| > 34 | 207,771 (19.1%) | 700,658 (18.0%) |
| Unknown/Implausible | 126 (0.0%) | 198 (0.0%) |
| Race/Ethnicity |  |  |
| Black, non-Hispanic | 146,572 (13.5%) | 413,508 (10.6%) |
| White, non-Hispanic | 385,096 (35.5%) | 1,401,767 (35.9%) |
| Hispanic | 379,973 (35.0%) | 1,413,141 (36.2%) |
| Other | 105,377 (9.7%) | 421,808 (10.8%) |
| Unknown | 68,144 (6.3%) | 250,961 (6.4%) |
| Education |  |  |
| Less than high school | 216,143 (19.9%) | 712,347 (18.3%) |
| High school/GED | 287,467 (26.5%) | 984,772 (25.2%) |
| More than high school | 545,330 (50.3%) | 2,088,829 (53.5%) |
| Unknown | 36,222 (3.3%) | 115,237 (3.0%) |

*PTB, preterm birth; ETB, early-term birth.*

**Table 2.**
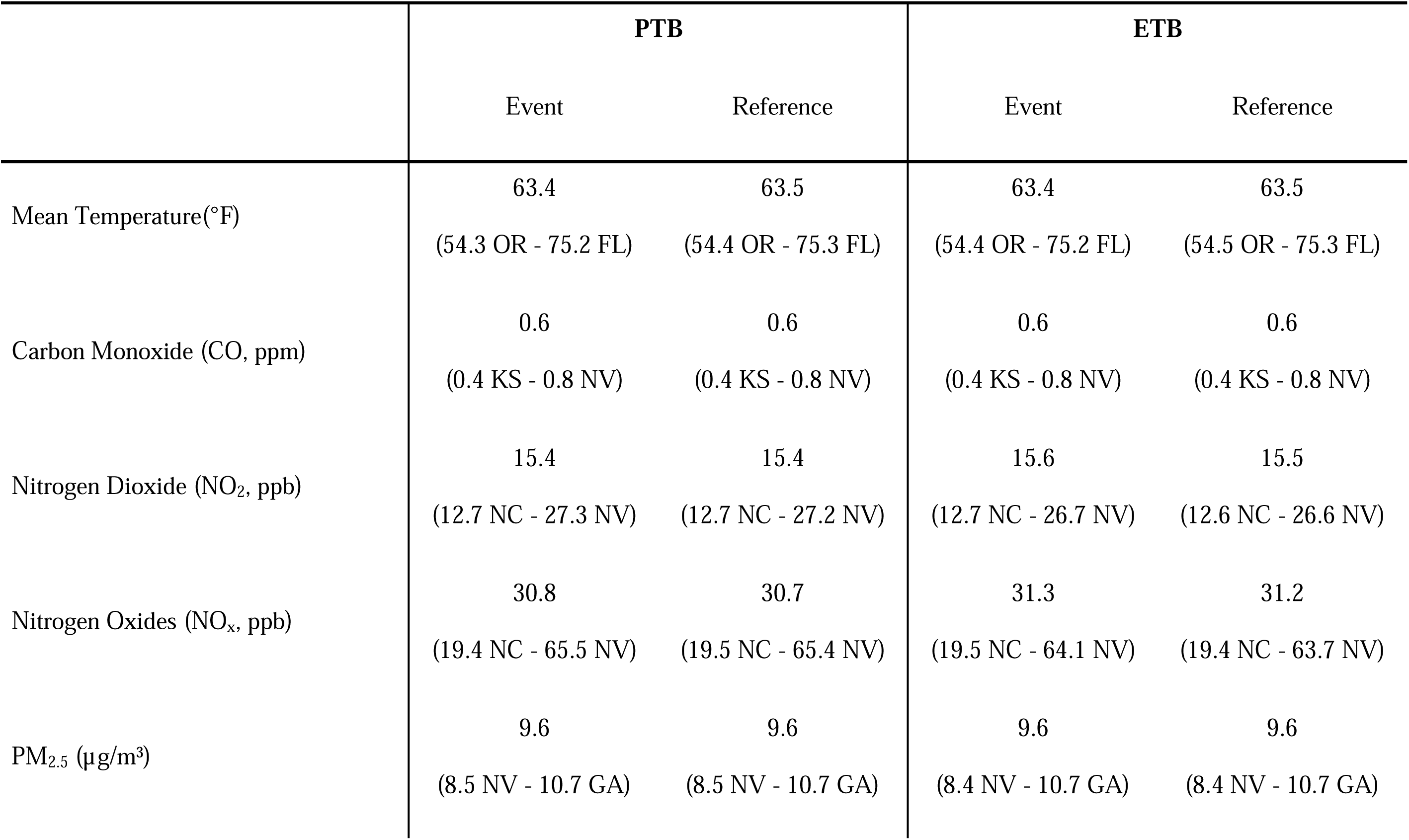
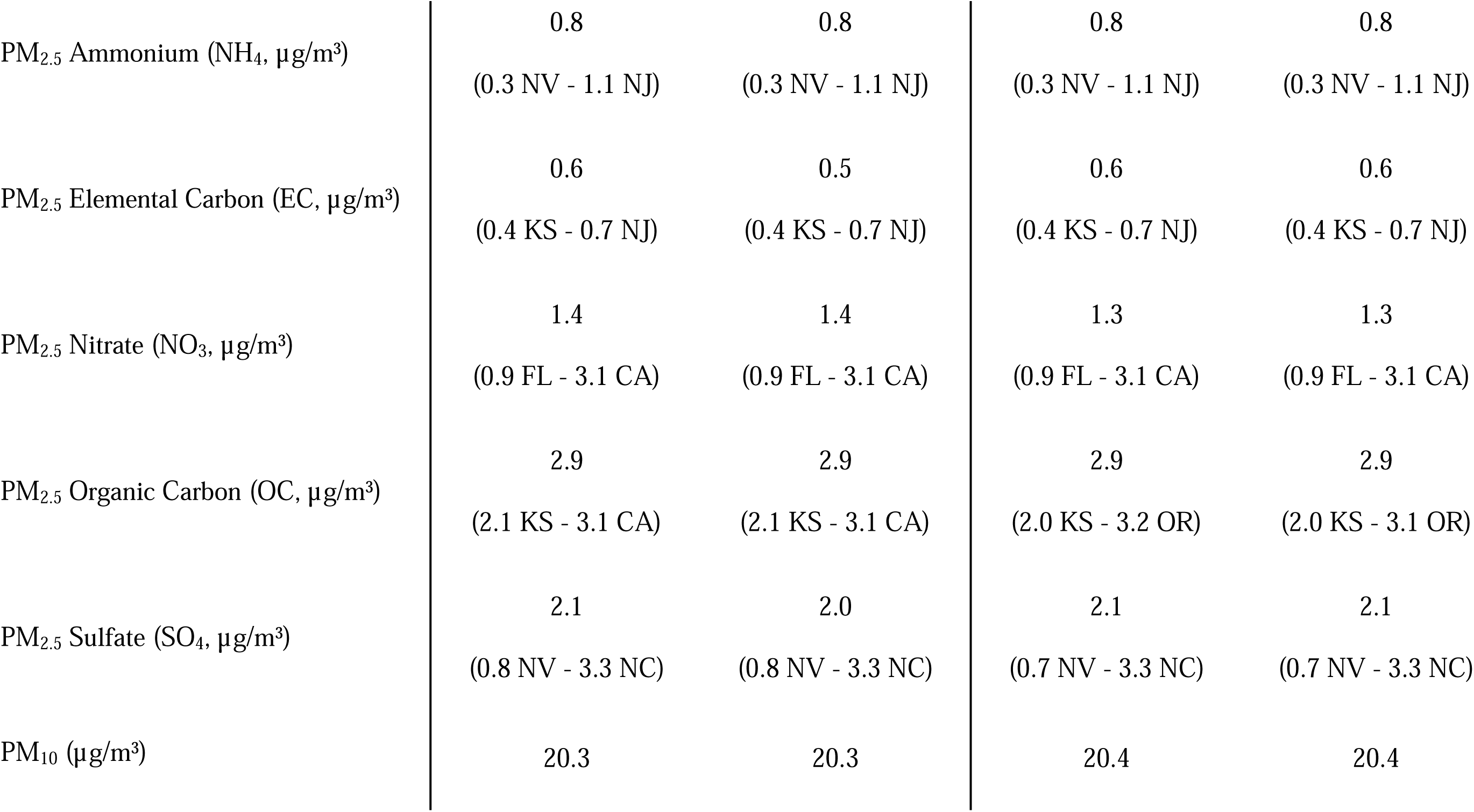
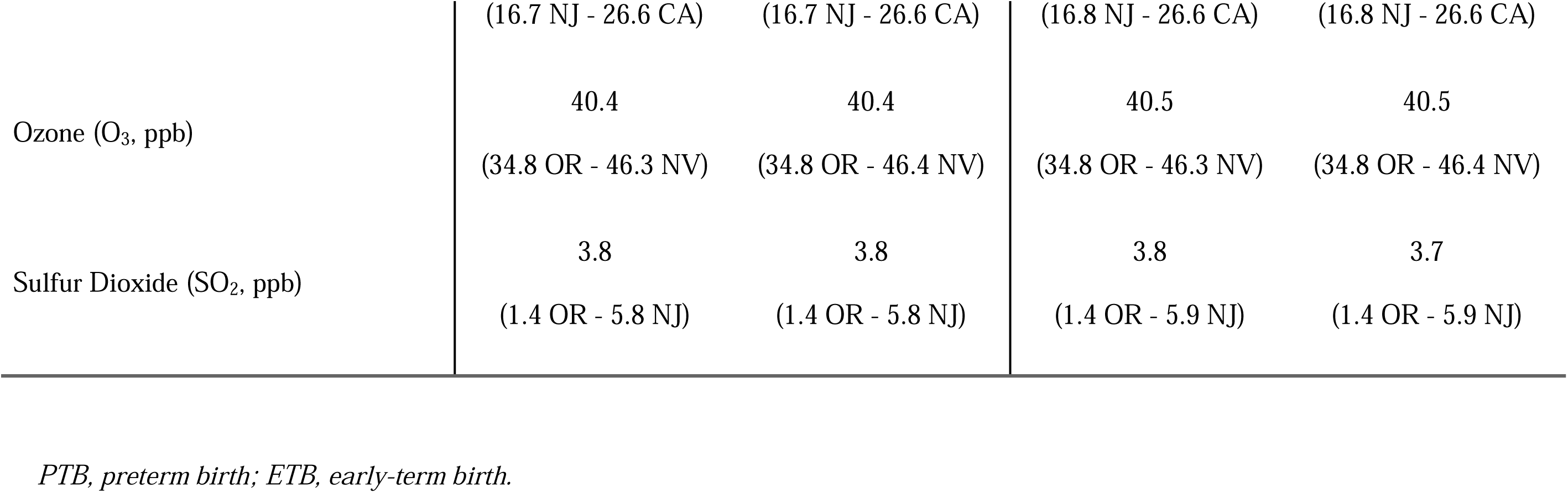
Median and range of average daily temperature and air pollution concentrations by event and reference days across states.

Figure 1 shows the pooled associations between short-term air pollution exposures and PTB or ETB. Criterion particulate matter and gaseous pollutants were found to be associated with ETB. The 0-2 lag cumulative exposure showed stronger associations compared to same-day (lag 0) exposure. Per IQR increase in exposure, NO_2_ (OR: 1.0023, 95% CI:1.0010, 1.0037) and PM_2.5_ (OR: 1.0022, 95% CI: 1.0006, 1.0038) showed the strongest 0-2 lag associations. Major constituents of PM_2.5_, except nitrate, were also found to be consistently associated with ETB. Per IQR increase in exposure, the strongest 0-2 day lag associations were with organic carbon (OR: 1.0026, 95% CI: 1.0013, 1.0039) and elemental carbon (OR: 1.0025, 95% CI: 1.0014, 1.0035). Associations for PTB were mostly null. Ozone was not associated with either ETB or PTB. All numerical estimates are provided in Online Supplemental Materials.

**Figure 1.**
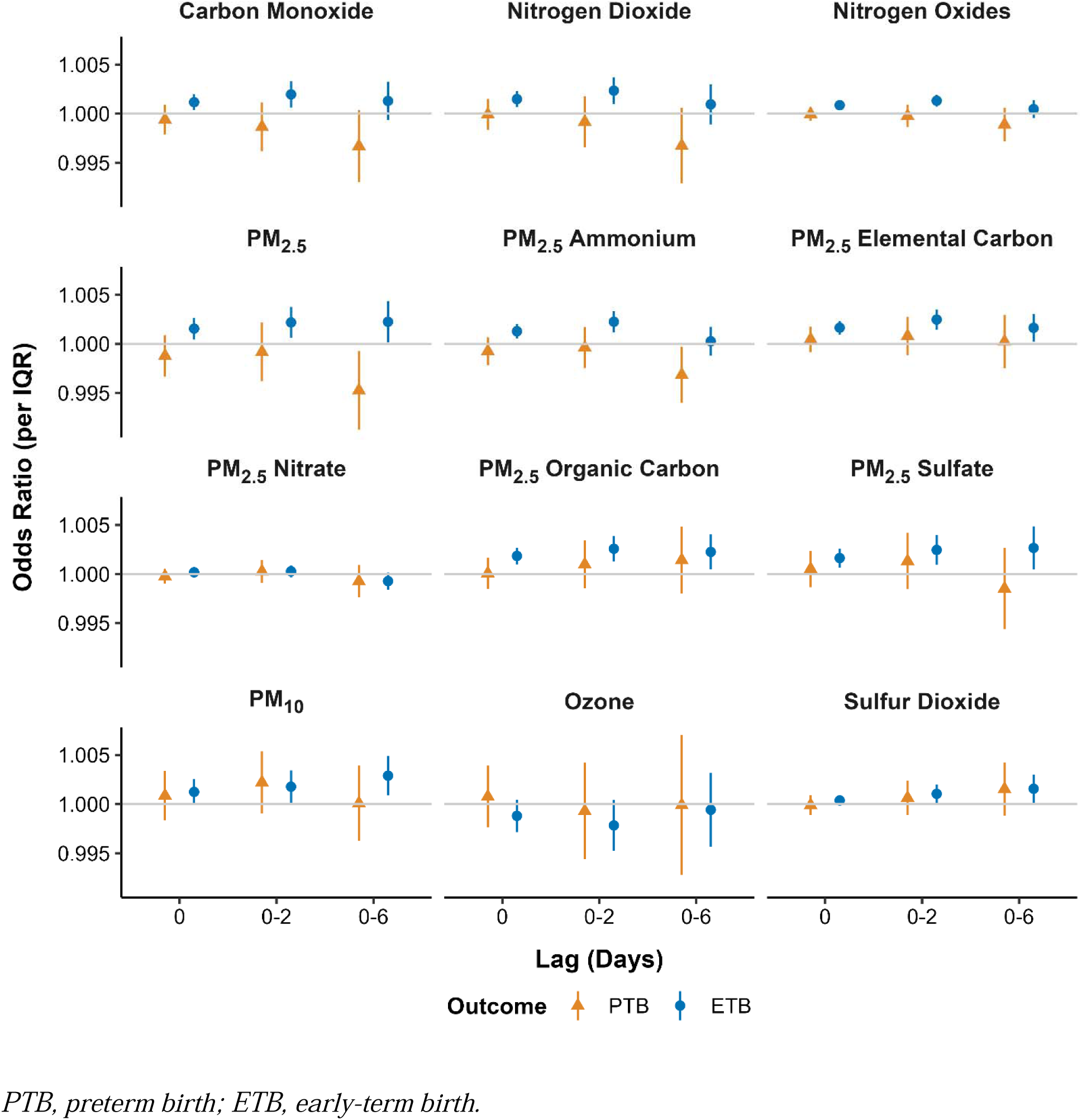
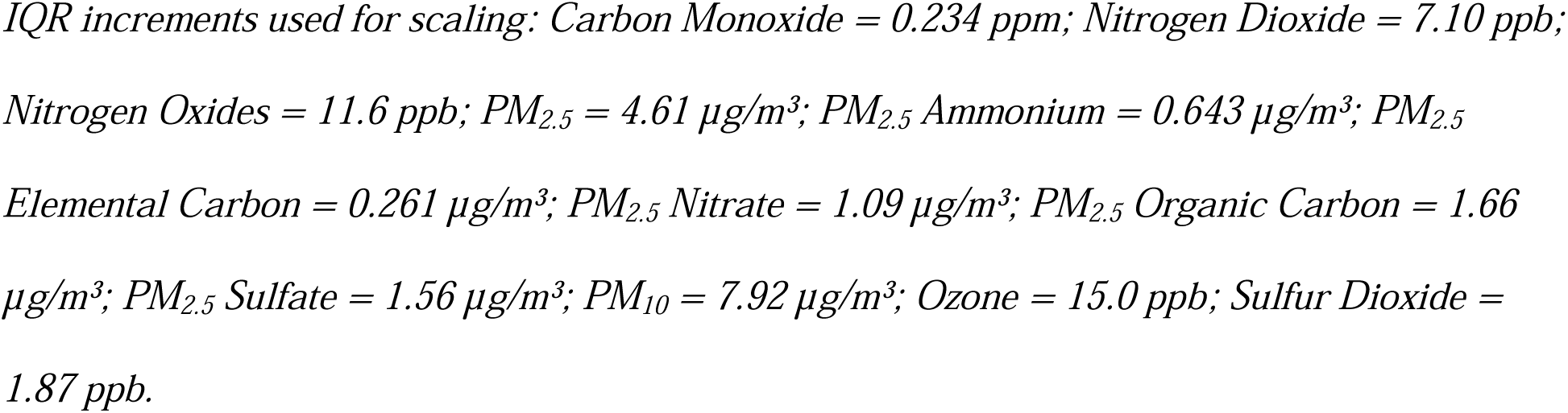
Association between ambient air pollution exposure and risk of preterm and early-term births. Odds ratios (OR) and 95% confidence intervals (CI) are expressed per interquartile range (IQR) increase in pollutant concentration. Results are shown for single-day (Lag 0) and cumulative (Lag 0-2, Lag 0-6) exposure windows. *PTB, preterm birth; ETB, early-term birth.* *IQR increments used for scaling: Carbon Monoxide = 0.234 ppm; Nitrogen Dioxide = 7.10 ppb; Nitrogen Oxides = 11.6 ppb; PM_2.5_ = 4.61 µg/m³; PM_2.5_ Ammonium = 0.643 µg/m³; PM_2.5_ Elemental Carbon = 0.261 µg/m³; PM_2.5_ Nitrate = 1.09 µg/m³; PM_2.5_ Organic Carbon = 1.66 µg/m³; PM_2.5_ Sulfate = 1.56 µg/m³; PM_10_ = 7.92 µg/m³; Ozone = 15.0 ppb; Sulfur Dioxide = 1.87 ppb*.

State-specific associations for the 0-2 lag exposure are given in eFigure 1. ETB was associated most consistently with NO_2_, PM_2.5_ total mass, PM_2.5_ elemental carbon, PM_2.5_ organic carbon, PM_2.5_ ammonium, and PM_2.5_ sulfate, with multiple states showing positive or null associations. North Carolina is the only state with a positive association between ETB and ozone (OR: 1.0121, 95% CI: 1.0032, 1.0211). Georgia is the only state with a positive association between ETB and PM_2.5_ nitrate (OR: 1.0100, 95% CI: 1.0046, 1.0155). State-specific associations for PTB were mostly null.

Figure 2 gives the associations between lag 0-2 air pollution exposure and PTB or ETB within each maternal age group. Positive associations with ETB were generally observed across all age groups, especially NO_2_, NO_x_, elemental carbon, and organic carbon. Notably, for PM_2.5_ and SO_2_, associations with ETB were null among those above age 34. When stratified by maternal race/ethnicity (eFigure 2), we found some evidence that associations between air pollution and ETB were stronger among Blacks (e.g., for PM_2.5_ OR: 1.0072, 95% CI: 1.0026, 1.0119) and Whites (PM_2.5_ OR 1.0029, 95% CI: 1.0002, 1.0057) compared to Hispanics (PM_2.5_ OR: 1.0004, 95% CI: 0.9979, 1.0029). When stratified by maternal education (eFigure 3), associations between air pollution and ETB were weaker among those with less than high school education for some pollutants. For PTB, the associations remained close to null across subgroups, with no clear evidence of effect modification by maternal age, race/ethnicity, or education (Figure 2, eFigure 2-3).

**Figure 2.**
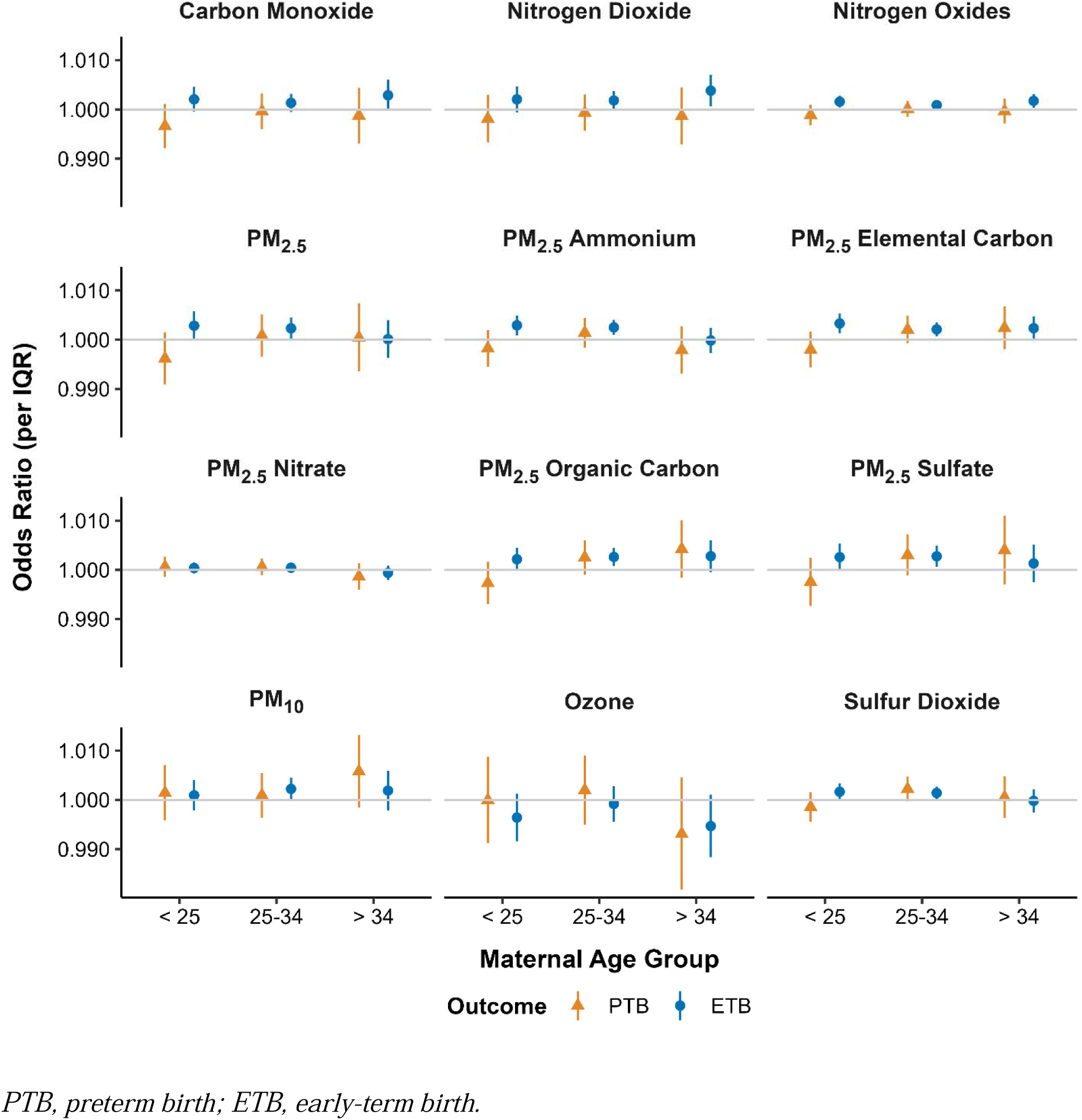
Effect modification of the association between cumulative air pollution exposure (lag 0-2) and birth outcomes by maternal age. Estimates represent the cumulative odds ratio (Lag 0-2) per IQR increase in pollutant concentration, stratified by maternal age group (<25, 25- 34, and > 34 years). *PTB, preterm birth; ETB, early-term birth*.

Extending the exposure window from 0-6 days to 0-14 days generally resulted in similar lag 0, lag 0-2, and lag 0-6 associations (eFigure 4). However, we also found that PTB was associated with 0-14 lag exposure for elemental carbon (OR: 1.0046, 95% CI: 1.0006, 1.0086) and organic carbon (OR: 1.0055, 95% CI: 1.0004, 1.0107). Our estimated associations are robust against increasing the degrees of freedom for the spline model for seasonality (eFigure 5).

## 4. Discussion

In this large multi-state study of nearly 5 million births across eight states of the United States, we found that short-term exposure to several ambient air pollutants was associated with a modest but consistent increase in the odds of ETB, whereas associations with PTB were largely null. The associations for exposures in the days immediately preceding delivery supporting the hypothesis that acute increases in air pollution may act as triggers of delivery . In contrast, we observed little evidence that short-term air pollution exposure substantially increases the risk of PTB. Overall, our findings suggest that acute air pollution exposure may influence gestational timing by modestly advancing delivery, rather than precipitating PTB. While ETB is less severe than PTB, ETB is a strong predictor of subsequent infant morbidity.

Several studies have reported associations between acute exposure to air pollutants in the days preceding delivery and increased risk of PTB, particularly for ozone and, in some cases, particulate matter. For example, a large multi-city time-to-event analysis in Canada observed increased hazards of PTB birth associated with ozone exposure within 0–3 days prior to delivery,^12^ and a nationwide case-crossover study in Sweden reported positive associations between short-term ozone exposure and PTB.^11^ Additional studies have identified short-term effects of particulate matter and gaseous pollutants across various lag structuresr.^20,21^ However, our results did not show consistent associations with PTB, but instead demonstrated more robust and consistent associations with ETB. As a result, studies focusing exclusively on PTB may underestimate the impact of acute environmental exposures on gestational duration. This interpretation is consistent with emerging evidence from studies of other acute environmental stressors, such as temperature extremes, which have reported stronger and more consistent associations with ETB than with PTB.^22^

The stronger and more consistent associations observed for ETB may be explained by ETB being a more sensitive and proximal outcome for detecting short-term environmental triggers. Compared with PTB, which is relatively rare and often driven by more severe or chronic pathological processes, ETB may be more responsive to transient physiological perturbations. Acute exposure to air pollutants has been linked to systemic inflammation, oxidative stress, and alterations in endocrine signaling and placental function, all of which could promote uterine activity or cervical ripening and thereby modestly advance the timing of delivery.^23,24^ These mechanisms are biologically plausible for inducing small shifts in gestational duration but may be insufficient to trigger earlier delivery into the preterm range. In addition, PTB is a heterogeneous outcome encompassing distinct etiologic pathways, including spontaneous and medically indicated subtypes, which may dilute associations when analyzed as a single endpoint.^25^ Together, these considerations highlight the importance of examining the full distribution of gestational age when evaluating the effects of short-term air pollution.

The observed associations across multiple pollutants, including PM_2.5_, PM_10_, NO_2_, and major PM_2.5_ constituents such as elemental carbon and organic carbon, further suggest that source-specific components of air pollution may play an important role in influencing gestational timing. Elemental carbon is commonly used as a marker of traffic-related emissions, whereas organic carbon can reflect a broader range of combustion sources, including biomass burning and wildfire smoke. Growing evidence indicates that these sources may differ in toxicity and health impacts. For example, wildfire smoke has been identified as an increasingly important contributor to ambient PM_2.5_ in the United States and has been associated with increased risk of PTB in large population-based studies.^15^ Traffic-related air pollution remains a major urban exposure of concern, although evidence linking it to PTB is limited. In our study, the consistency of associations across multiple pollutants and PM_2.5_ constituents supports the hypothesis that combustion-related pollution mixtures may be particularly relevant for short-term effects. At the same time, the high correlation among pollutants limits the ability to disentangle independent effects of specific sources, and the observed associations may reflect the combined influence of complex pollutant mixtures rather than single pollutants in isolation.^11,12^

We also observed evidence of effect modification across several population subgroups, suggesting that susceptibility to short-term air pollution exposure may not be uniform. Stronger associations were observed among non-Hispanic Black and non-Hispanic White mothers, as well as among younger maternal age groups, although patterns were not consistent across all pollutants. These differences may reflect a combination of biological susceptibility, differential exposure patterns, and underlying social and environmental determinants of health. For example, structural inequities and residential segregation may result in higher exposure to traffic-related or combustion-related pollution among certain populations, while differences in access to healthcare, baseline health status, or co-exposures such as psychosocial stress could further modify vulnerability. Prior studies have similarly reported heterogeneous associations across sociodemographic subgroups, including stronger effects among populations with higher baseline risk or environmental burden.^7,8,26^ At the same time, some subgroup findings may be influenced by exposure misclassification, resulting in attenuated associations.

This study has several notable strengths. First, it leverages a very large sample size of nearly 5 million births across eight states in the United States, providing substantial statistical power to detect small effect sizes. Second, the inclusion of geographically and demographically diverse populations enhances the generalizability of the findings. Third, the time-stratified case-crossover design effectively controls for time-invariant individual-level confounders and reduces bias from seasonal and long-term trends. Fourth, we examined a comprehensive set of twelve pollutants, including major PM_2.5_ constituents, allowing for a more detailed assessment of pollutant mixtures and potential source-specific effects.

This study also has several limitations. First, exposure misclassification is possible because air pollution levels were derived from a data product at 12km spatial resolution, which cannot adequately resolve intra-urban variation. However, our analyses mostly utilize temporal variations in exposure. Area-level exposure estimates also do not reflect individual-level exposure, particularly given variation in time–activity patterns and indoor infiltration. For example, studies have shown that factors such as housing characteristics can influence the degree to which outdoor air pollution penetrates indoor environments, potentially leading to measurement error.^16^ Second, we were unable to account for individual-level factors that may vary over short time scales, such as acute infections, psychosocial stress, or physical activity, which could contribute to residual confounding even after adjusting for ambient temperature and temporal trends The use of area level ambient air pollution may minimize impacts individual-level confounders.^27^ Third, although the case-crossover design controls for time-invariant confounders, residual confounding by time-varying factors cannot be fully ruled out. Fourth, we did not differentiate between subtypes of PTB, such as spontaneous and medically indicated deliveries, which may have distinct etiologies and sensitivities to environmental exposures.^25^

Despite the modest effect sizes observed, our findings have important public health implications. ETB is substantially more common than PTB and is increasingly recognized as a clinically meaningful outcome. Therefore, even small shifts in gestational timing at the population level could translate into a considerable burden of adverse outcomes. The identification of short-term exposure windows immediately preceding delivery also suggests opportunities for targeted interventions, such as real-time air quality advisories, behavioral recommendations to reduce exposure during high pollution days, and clinical guidance for pregnant individuals nearing term. In addition, our results underscore the potential benefits of policies aimed at reducing emissions from major sources of air pollution, including traffic and combustion-related activities.

In conclusion, this large multi-state study provides evidence that short-term exposure to ambient air pollution is associated with modest increases in the risk of ETB, but not for PTB. These findings indicate that acute environmental exposures may act as triggers that advance delivery timing. By incorporating a comprehensive set of pollutants and focusing on short-term exposure windows, this study fills an important knowledge gap and highlights the importance of considering both exposure timing and the full distribution of gestational age. Future studies should further investigate source-specific exposures and refine exposure assessment to better understand underlying mechanisms and identify opportunities for prevention.

## Data Availability

Birth record data must be obtained directly from state health departments.
Meteorology data are available at: https://rda.ucar.edu/datasets/d314008/. Air pollution data will be made available on request.

---

**eFigure 1.**
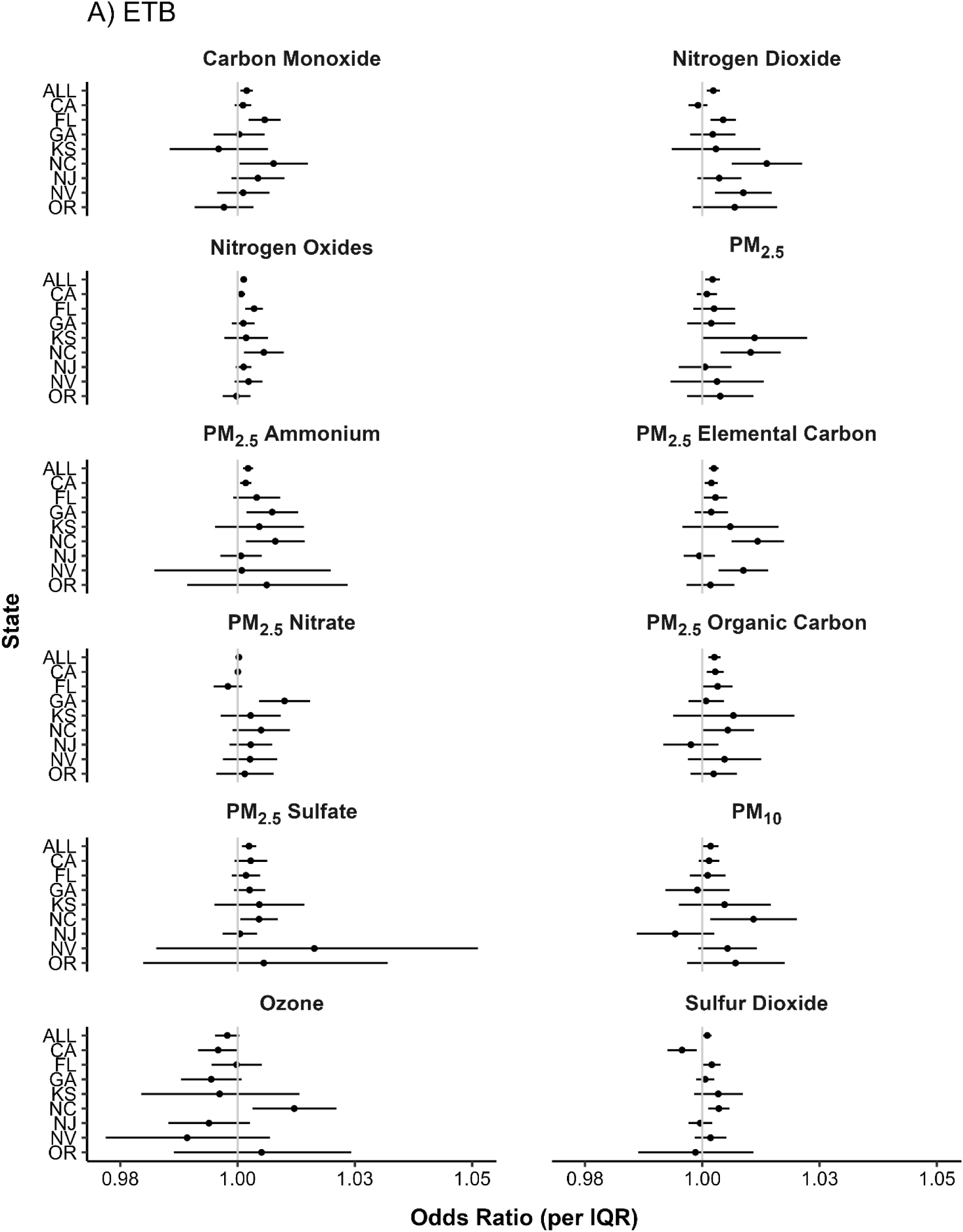
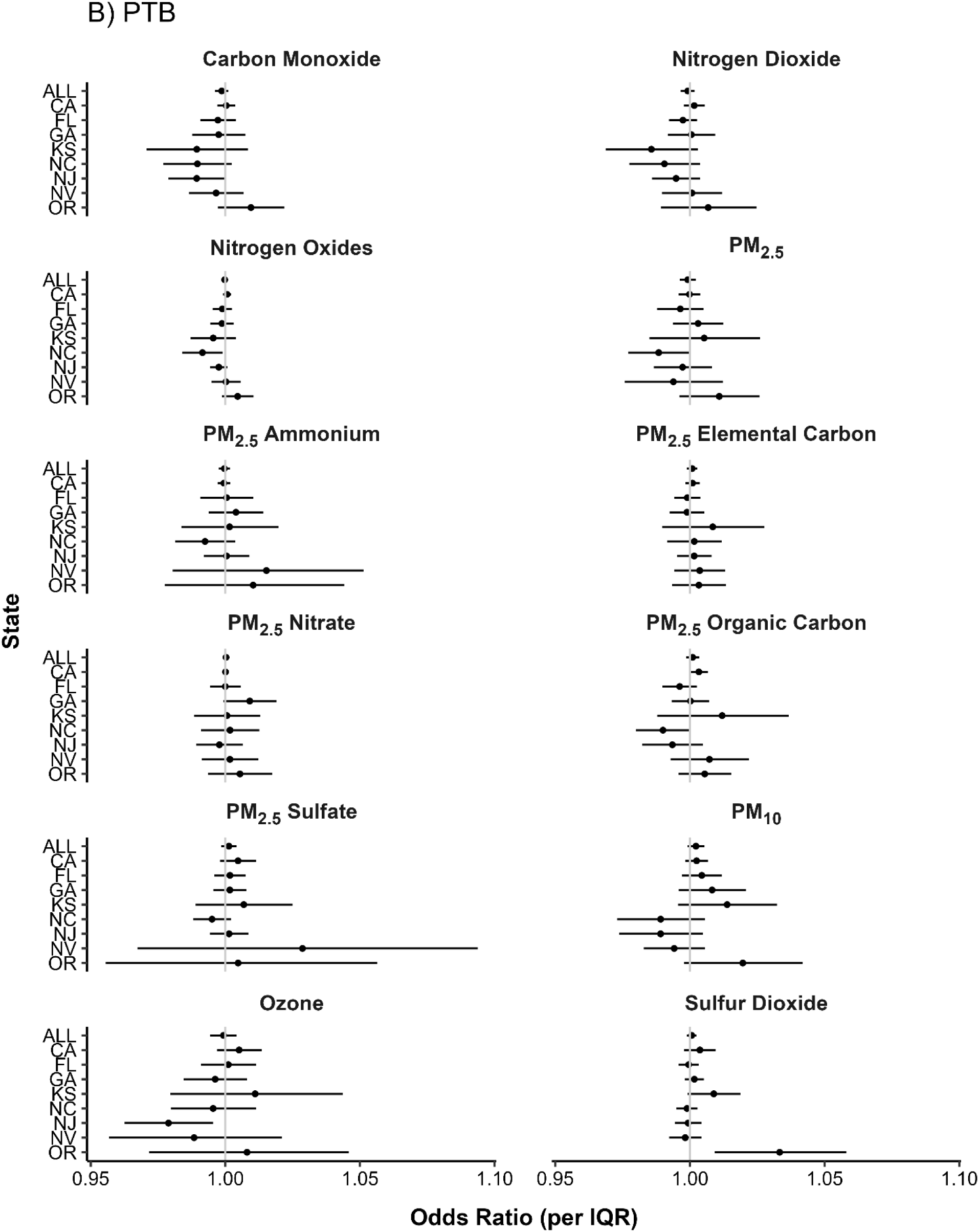
State-specific associations between cumulative air pollution exposure (Lag 0-2) and (A) Early-term Birth (ETB) or (B) Preterm Birth (PTB).

**eFigure 2.**
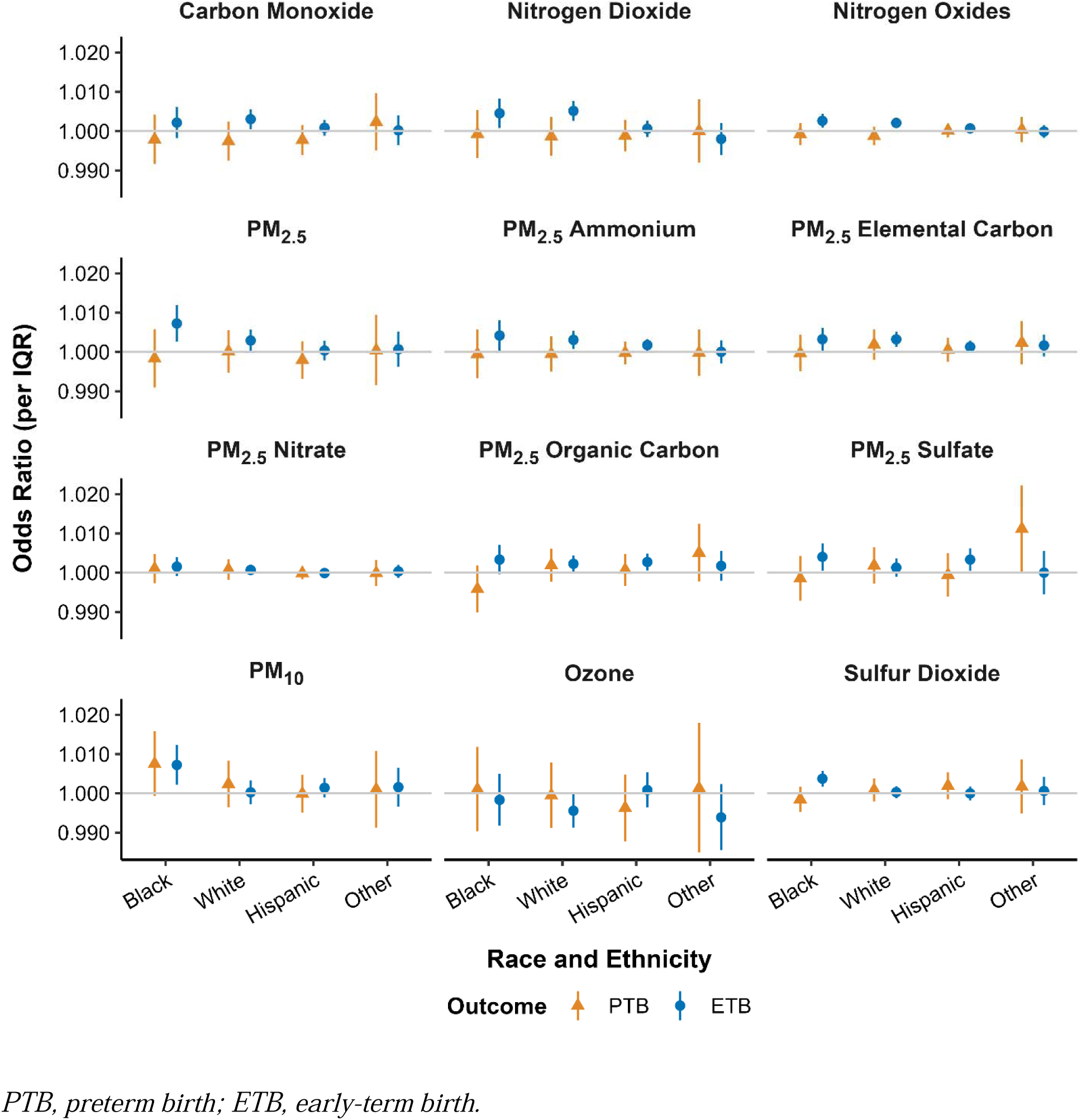
Effect modification of the association between cumulative air pollution exposure (Lag 0-2) and birth outcomes by maternal race and ethnicity. Estimates represent the cumulative odds ratio (Lag 0-2) per IQR increase in pollutant concentration, stratified by race andethnicity. *PTB, preterm birth; ETB, early-term birth*.

**eFigure 3.**
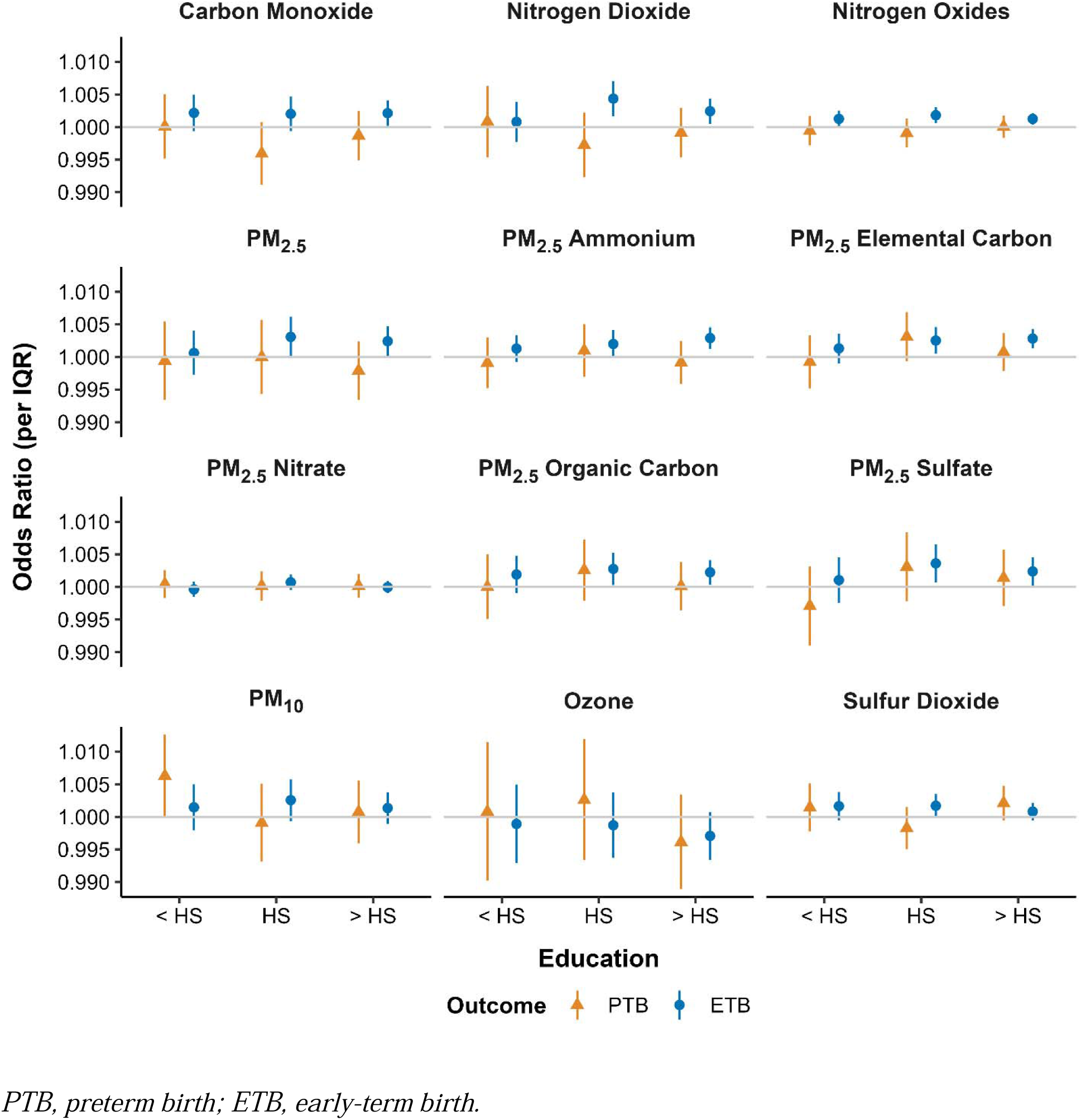
Effect modification of the association between cumulative air pollution exposure (Lag 0-2) and birth outcomes by maternal education level. Estimates represent the cumulative odds ratio (Lag 0-2) per IQR increase in pollutant concentration, stratified by education level: less than high school (<HS), high school (HS), and more than high school (>HS). *PTB, preterm birth; ETB, early-term birth*.

**eFigure 4.**
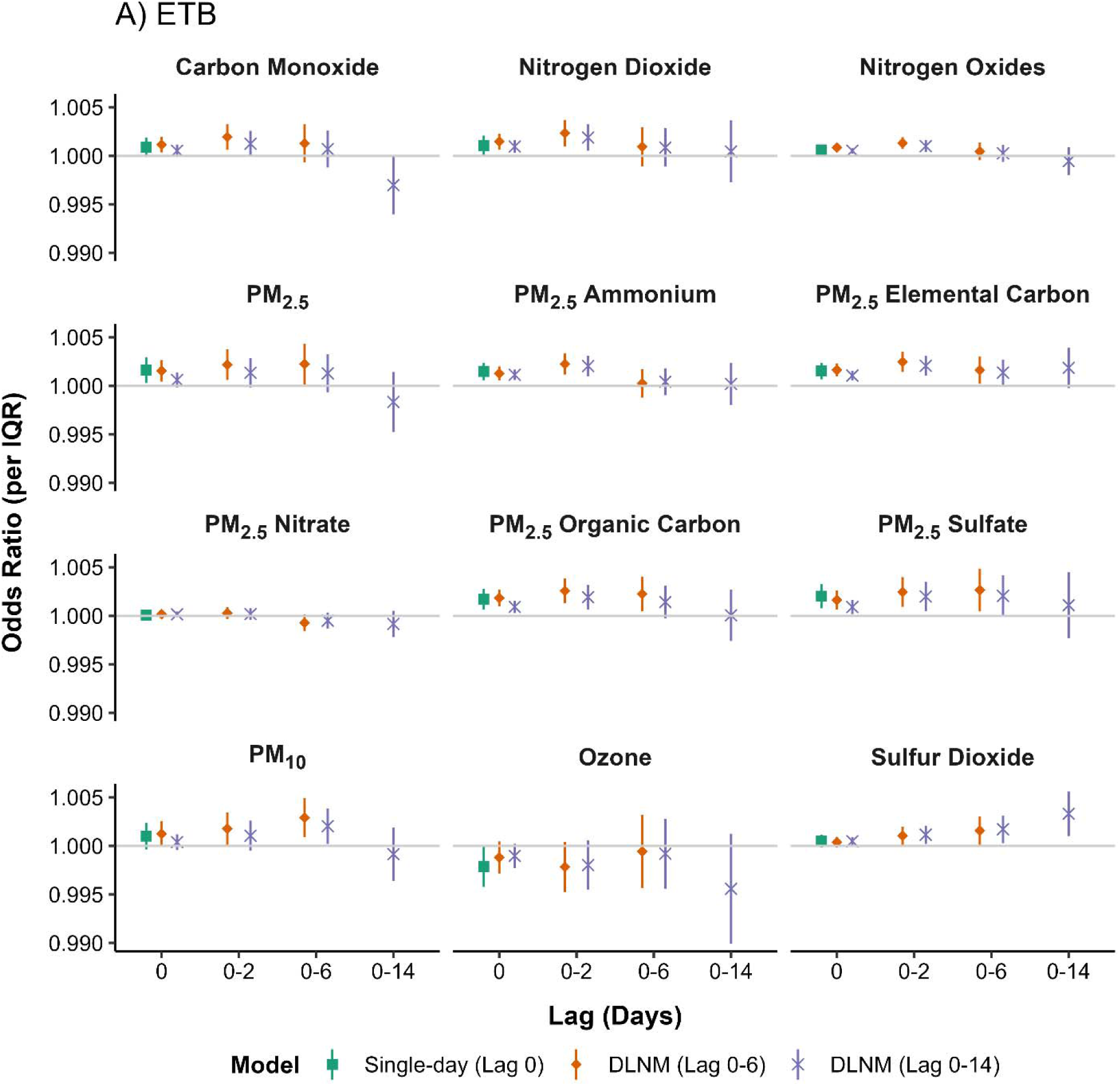
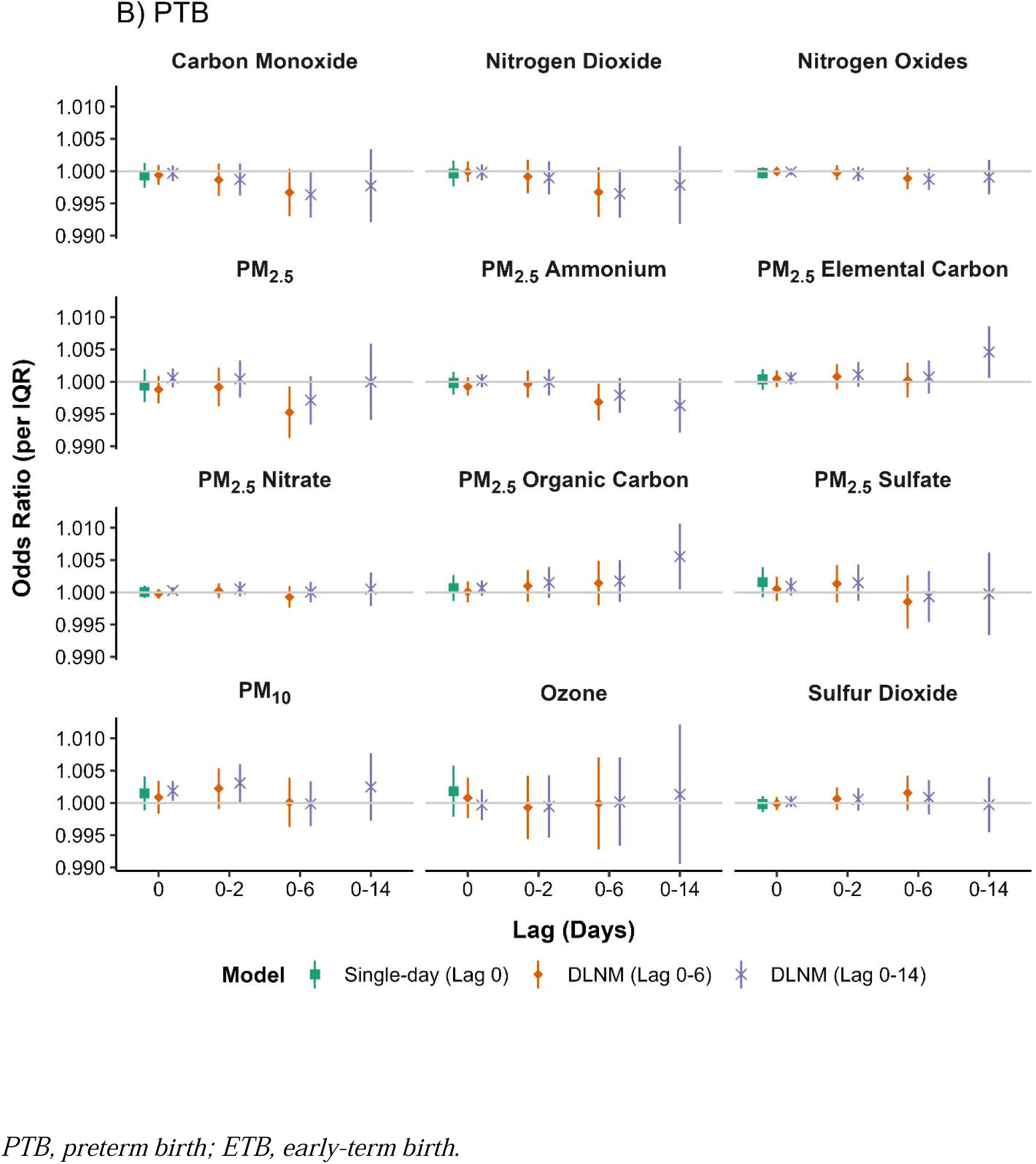
Sensitivity of results to model specification for (A) Early-term Birth and (B) Preterm Birth. Comparison of odds ratios derived from three different model specifications: a single-day exposure model (Lag 0) and distributed lag models covering 0–6 days (the primary model) and 0–14 days. *PTB, preterm birth; ETB, early-term birth*.

**eFigure 5.**
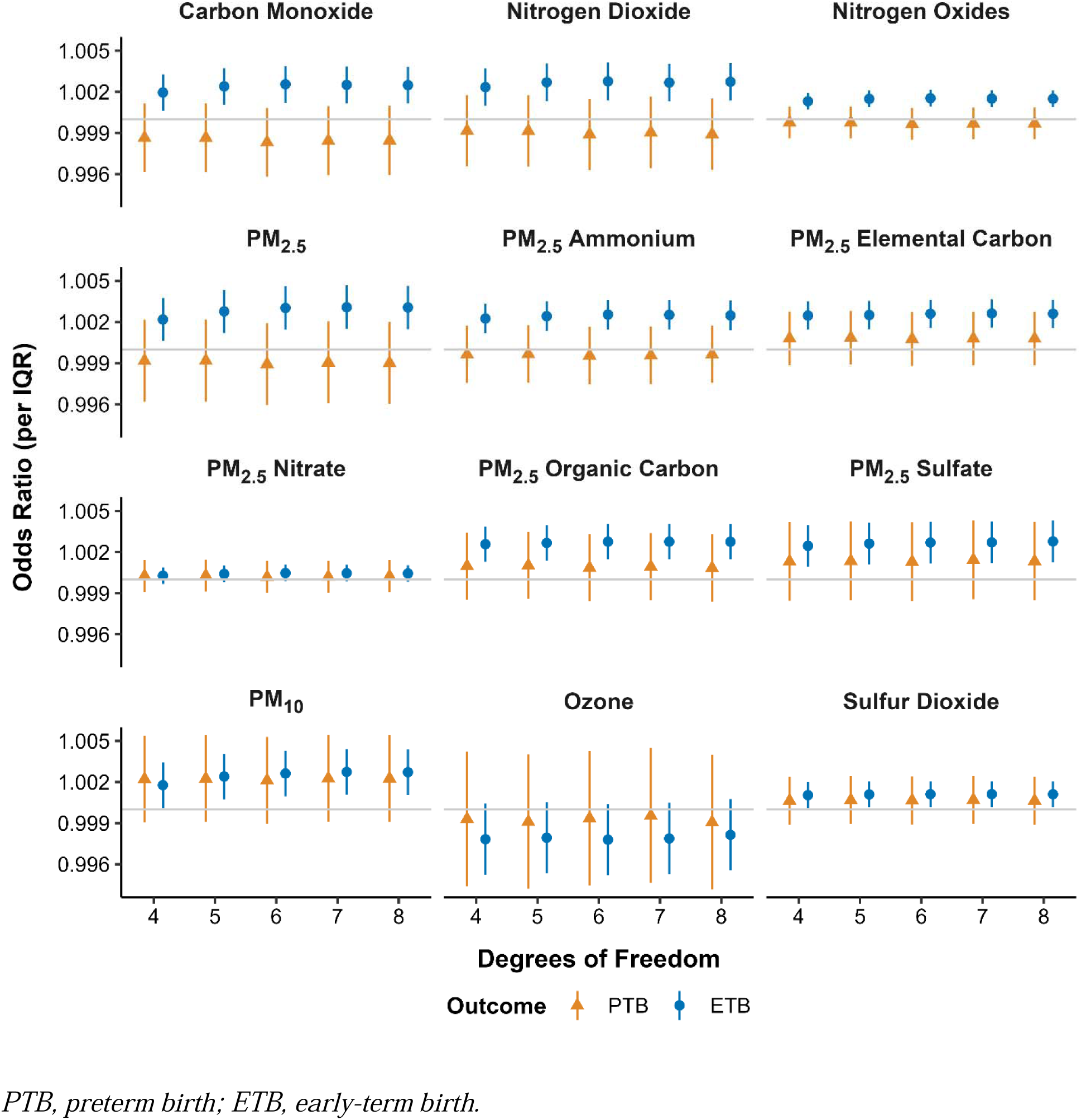
Sensitivity of the association between cumulative air pollution exposure (Lag 0- 2) and birth outcomes to the degrees of freedom (df) used for seasonality control. Results are shown for seasonality splines with df ranging from 4 to 8 per year. *PTB, preterm birth; ETB, early-term birth*.

**eTable 1.** Number and percent of preterm and early-term cases between 2005-2017 (NC: 2005-2015) by state.

| State | PTB n (%) | ETB n (%) | Total n (%) |
| --- | --- | --- | --- |
| CA | 447,594 (41.2%) | 1,639,545 (42.0%) | 2,087,139 (41.9%) |
| FL | 208,794 (19.2%) | 777,846 (19.9%) | 986,640 (19.8%) |
| GA | 142,010 (13.1%) | 479,909 (12.3%) | 621,919 (12.5%) |
| KS | 33,553 (3.1%) | 124,900 (3.2%) | 158,453 (3.2%) |
| NC | 99,050 (9.1%) | 319,333 (8.2%) | 418,383 (8.4%) |
| NJ | 85,097 (7.8%) | 311,170 (8.0%) | 396,267 (7.9%) |
| NV | 37,092 (3.4%) | 126,126 (3.2%) | 163,218 (3.3%) |
| OR | 31,972 (2.9%) | 122,356 (3.1%) | 154,328 (3.1%) |
| Total | 1,085,162 | 3,901,185 | 4,986,347 |

*PTB, preterm birth; ETB, early-term birth.*

## Footnotes

^1^Maternal ages falling outside the range 10-55 are considered implausible and excluded from the effect modification study.

